# Burden and trends of endoscopically unresectable polyps requiring surgical intervention: A systematic review and meta-analysis

**DOI:** 10.64898/2026.09.15.26363122

**Authors:** M. Mustafa Arif, Muhammad Tariq Bakhshi, Maryam Sherwani, Alan Kuntz, Nithesh Kumar, Alejandro Chara, Aimal Khan

## Abstract

**Background:** Colorectal polyps judged endoscopically unresectable are conventionally referred for colectomy, carrying substantial morbidity for a lesion that is usually benign. Advanced endoscopic techniques have made many such lesions resectable, but the burden and temporal trends of surgery remain unclear.

**Methods:** PubMed, Scopus, Embase, Web of Science, and Cochrane CENTRAL were searched from inception to 1 September 2026 for studies reporting surgery for endoscopically unresectable polyps against a population or health-system denominator. Estimates were assigned a priori to four denominator classes: population at risk (N1), colorectal resections (N2), polyps detected (N3), or colonoscopies/screening episodes (N4). Estimates were analysed within class and never combined across classes. Within denominator sub-definitions containing at least three independent data sources, proportions were pooled using random-effects models on the logit scale with restricted maximum likelihood estimation of between-study variance, Hartung-Knapp adjusted confidence intervals and 95% prediction intervals. Risk of bias was assessed using the JBI checklist for prevalence studies and certainty using an adapted GRADE approach.

**Results:** Eighteen studies contributed 122 burden estimates; 105 of 130 full texts were excluded for lacking a population or health-system denominator. Seventy-five distinct denominator definitions were identified. Estimates ranged from 1.09%–38.38% of colorectal resections and 0.09%–52.38% of polyps detected. Seven studies reported one numerator against multiple denominators, producing within-study differences of up to 88.3-fold. When denominators matched, estimates converged: 12 values from two national datasets spanned only 0.20%–0.37% of unselected colonoscopy volume. Unresectability criteria were reported in 7 of 17 assessable studies; only one satisfied all five reporting items. Three sub-definitions contained enough independent sources to pool, and every pooled estimate carried a wide prediction interval: 23.79% (95% CI 12.27-41.06; 95% prediction interval 3.22-74.55) of resections for colorectal neoplasia, 1.15% (0.31-4.21; 0.02-37.19) of all detected polyps and 17.11% (4.36-48.30; 0.31-93.22) of large or complex lesions. Restricting estimates to a single denominator sub-definition reduced between-study variance by 53% among polyp-based denominators.

**Conclusions:** The burden of surgery for endoscopically unresectable colorectal polyps cannot presently be expressed as a single figure. Published disagreement is primarily definitional, with estimates converging when denominators match. Surgical volume appears to have peaked around 2012–2014 and subsequently declined. A minimum reporting standard is needed to measure this burden reliably.

## Introduction

Colorectal polyps are commonly seen on lower gastrointestinal endoscopy, with reported prevalence ranging from 20-38% across studies. A large proportion of polyps are now resected endoscopically with newer endoscopic techniques**^(1–3)^**. However, some lesions may be considered to be endoscopically unresectable polyps (EUPs) because of size, morphology, location, or concern for malignant characteristics**^(4,5)^**. Conventionally, patients with polyps labelled as EUPs were referred for operative intervention**^(6)^**. Although surgery allows for definitive treatment**^(4)^**, colorectal resection is associated with significant morbidity compared to endoscopic management**^(7)^**. Therefore, EUPs represent a significant problem in clinical practice at the interface between endoscopic and surgical management.

Evolving endoscopic techniques have significantly changed the management of EUPs, enabling polyps previously labelled unresectable to now be removed without the added morbidity of surgery**^(8)^**. As the role of advanced endoscopic management has expanded, the prevalence of surgery for EUPs and the characteristics of patients requiring surgery may also be changing. Although recent studies have demonstrated a decline in surgical resections for nonmalignant polyps**^(9,10)^**, the overall burden, temporal trends, and patient characteristics remain incompletely understood. To date, no systematic review has collated the available evidence for this patient population.

To address this gap, we conducted a systematic review to synthesize the available literature on the burden and temporal trends of EUPs managed surgically. Quantifying this burden as an aggregate incidence and as a proportion of overall colorectal surgical volume is essential to understand if advanced endoscopy results in fewer operations. We hypothesize that the incidence of EUPs requiring surgical management has declined over time, along with the expansion of recent advanced endoscopy techniques.

## Methodology

### Study Design and Protocol Registration

This systematic review followed the Preferred Reporting Items for Systematic Reviews and Meta-Analyses (PRISMA) 2020 statement(^11^). The protocol was pre-registered on the International Prospective Register of Systematic Reviews (PROSPERO) with registration ID: CRD420261494527 for transparency and methodological consistency. The completed PRISMA 2020 checklist is provided in **Appendix 1**.

### Eligibility Criteria

This systematic review was guided by a predefined PECO (Population, Exposure, Comparison, Outcome) framework. The population comprised of adults aged 18 years or older undergoing lower gastrointestinal endoscopy in whom one or more colorectal polyps were detected. The exposure of interest was the designation of a colorectal polyp reported by the study as endoscopically unresectable, non-resectable, unsuitable for endoscopic removal, or otherwise considered to require surgical management. This was a single-arm review with no comparator group; where an included study also reported an endoscopically managed arm, those data were extracted only in order to define the denominator.

Eligible study designs were population-based and registry analyses, administrative database studies, prospective and retrospective cohort studies, case series, cross-sectional studies, and the relevant arms of comparative observational studies and randomized trials. Narrative reviews, editorials, guidelines, case reports, and animal or ex vivo studies were excluded, although the reference lists of reviews and guidelines were searched by hand. Studies confined to patients with inflammatory bowel disease-associated dysplasia, hereditary polyposis syndromes, or biopsy-proven invasive adenocarcinoma at the time of designation were also excluded, as the resectability decision in these populations is governed by different considerations.

### Outcomes

The primary outcome was the burden of surgery for EUPs, expressed as a proportion or rate against an explicitly reported denominator, alongside its temporal evolution. Due to the published literature quantifying this burden against several different types of denominators which are not interchangeable, every estimate was assigned a priori to one of four denominator classes: the population at risk (N1), expressed per 100,000 persons; all colorectal resections (N2); all polyps, or all complex polyps, detected (N3); and all colonoscopies or screening episodes (N4). Estimates were then analyzed within each denominator class and were not combined across different classes.

Secondary outcomes included the criteria and case-ascertainment methods used to designate a colorectal polyp endoscopically unresectable and also the completeness with which these were reported, and secondly, the temporal trends for surgeries performed for EUPs.

### Sources of Data and Search Strategy

Two independent authors conducted a systematic literature search across the PubMed (MEDLINE), Scopus, Embase, Web of Science, and Cochrane CENTRAL databases. The search encompassed all relevant studies from database inception through September 1st, 2026. Our strategy employed a combination of Medical Subject Headings (MeSH) and free-text terms specifically related to “colorectal polyps,” “endoscopically unresectable,” “colectomy rates,” and “difficult endoscopy”. The search string was further verified by a librarian to make sure there were no errors in the Boolean operators. A detailed description of the complete search strings is provided in **Appendix 2**.

### Study Selection Process

Following the systematic search, all identified citations were exported to Covidence**^(12)^**, where duplicate records were automatically removed. The remaining studies underwent initial screening of titles and abstracts, performed independently by two pairs of reviewers. Articles that potentially met the inclusion criteria, or those requiring further clarification, were retrieved for full-text evaluation. Any discrepancies during the screening or selection phases were resolved through discussion and consensus; if an agreement could not be reached, a third reviewer acted to resolve disagreements. Only studies published in English were considered for inclusion. No geographic restrictions were applied.

### Data Extraction

Data was independently extracted by two sets of reviewers using a standardized data extraction form, with discrepancies resolved by a third reviewer. Extracted parameters included the first author, year of publication, country, data source and source type, the sampling frame as described by the authors, the calendar years of observation, study design, whether an organized colorectal cancer screening program was operating, the number of patients and of lesions, funding, and publication type.

For case ascertainment, we extracted the case definition verbatim, any diagnostic or procedure codes used together with whether those codes had been validated, whether histological confirmation of benignity was required, how lesions found to be malignant on the resection specimen were handled, and whether emergency presentations, inflammatory bowel disease and polyposis syndromes were excluded.

For every reported estimate, we extracted the numerator, the denominator value, the definition of that denominator verbatim, its assigned denominator class, the observation period, the estimate as published with any confidence interval, and any reported trend statistic, direction, and P value. For the first secondary outcome we extracted five prespecified reporting items: whether any criteria for endoscopic unresectability were stated; whether those criteria were operationally precise; who applied the designation; at what point in the care pathway it was applied; and whether it was applied prospectively or ascertained retrospectively.

Every proportion was independently recomputed from the reported numerator and denominator before entry; discrepancies between the recomputed and the published value were recorded, and any figure derived by the review team rather than reported by the authors was labelled as such with the arithmetic stated. Where multiple included studies drew from overlapping years of the same administrative database or registry, only the study with the most complete or longest reported time span was retained for the primary burden/trend synthesis; overlapping studies were noted narratively but not treated as independent data points. Studies reporting data from a single calendar year or time point contributed to burden estimates only; studies reporting data spanning ≥2 discrete time points or a continuous multi-year period were eligible to contribute to the temporal trend synthesis.

### Risk of Bias and Publication Bias

The risk of bias in individual studies was initially assessed by two reviewers using the JBI Critical Appraisal Checklist for Prevalence Studies**^(13)^**. This assessment was then rechecked by a third reviewer. In addition to the checklist items, five supplementary items specific to administrative and registry data were applied to every study contributing a burden estimate: representativeness of the sampling frame, completeness of the reported code list, stability of the denominator across the observation period, handling of any coding revision falling within that period, and whether non-elective operations were excluded.

Publication bias was not formally evaluated, because no denominator class contained ten or more independent data sources, which is the minimum amount generally regarded as necessary for meaningful assessment of funnel plot asymmetry.

### Certainty of Evidence Assessment

Certainty of evidence was assessed using the adapted GRADE approach**^(14)^** for prevalence data by two independent reviewers.

### Data Synthesis and Statistical Analysis

Findings were synthesized within denominator class and, within class, between prespecified denominator sub-definition. Because the four classes address different questions, estimates were never combined across classes, and no overall summary estimate is presented. Within a sub-definition containing estimates from at least three independent data sources, proportions were pooled using a random-effects model fitted on the logit scale. The logit transformation was preferred to the Freeman-Tukey double arcsine transformation because back-transformation of the latter is unreliable when proportions approach zero, and many estimates in this review lie below 1%. Between-study variance was estimated by restricted maximum likelihood, confidence intervals used the Hartung-Knapp adjustment, and a continuity correction of 0.5 was applied. For each pooled estimate, we report the pooled proportion, its 95% confidence interval, the between-study variance, I-squared and a 95% prediction interval derived on t with k-2 degrees of freedom. Prediction intervals accompany every pooled proportion and are the primary measure of dispersion, because with heterogeneity of this magnitude a confidence interval around the mean conveys little about the range within which a future estimate would fall. Sub-definitions containing fewer than three independent data sources were not pooled and are reported descriptively. To avoid counting the same patients more than once, studies drawing on the same underlying data source were collapsed into a single unit and one estimate was retained per independent source per sub-definition, chosen mechanically as the estimate with the largest denominator. The contribution of sub-definition to between-study variance was quantified by comparing the variance estimated with all sources of a class combined against the source-weighted mean of the variances estimated within each sub-definition. This quantitative synthesis was not specified in the registered protocol, which anticipated narrative synthesis alone; it was added to characterize heterogeneity formally, and the pooled proportions are presented to describe dispersion rather than as recommended summary measures of burden.

### Organization of Outcomes

Outcomes were organized by denominator class, with each estimate plotted against the midpoint of its observation period. Separate panels were used for each denominator class. Temporal trends were displayed alongside one another but were not combined into a pooled slope. The available trend estimates were derived from a small number of overlapping national datasets using different denominators and observation periods.

### Recalculation and Summary of Estimates

When both a numerator and denominator were reported, the proportion was recomputed as numerator/denominator × 100, and the recomputed value was used for analysis. This ensured consistency across estimates.

Summary values are presented as observed ranges and medians. To avoid giving greater weight to sources reporting multiple annual estimates, medians were calculated in two stages. First, the median of estimates contributed by each independent data source was calculated, followed by the median across data sources. Medians were reported only within denominator sub-definitions and were not calculated at the broader denominator-class level, as this would combine estimates representing different underlying quantities.

Estimates for which the authors did not report both a numerator and denominator, estimates expressed as population rates, and estimates with an unresolved arithmetic discrepancy were displayed descriptively but excluded from quantitative comparison because their variance could not be calculated.

### Assessment of Inter-Study Variation

Pooling was performed only within a denominator sub-definition and never across denominator classes. The criteria used to designate a polyp as endoscopically unresectable, and the rules determining which lesions contributed to the numerator, varied substantially between studies; a proportion pooled across an entire class would therefore have represented different underlying quantities and would not have been interpretable. Pooled proportions are consequently presented within sub-definition only, and always with a prediction interval, so that the dispersion they summarize remains visible.

Because estimates within each denominator class did not represent a common underlying quantity, and several studies contributed repeated estimates from the same underlying data source, heterogeneity statistics supporting the pooled estimates are reported within sub-definition and after restriction to one estimate per independent source. Variation is described using observed ranges, source-level medians, between-study variance, I-squared and prediction intervals.

### Reporting Completeness and Study Characteristics

Completeness of case-definition reporting was summarized using the five prespecified reporting items. Patient and polyp characteristics were tabulated by study and observation period. The five reporting items were summed to generate a score out of five and categorized as complete (5/5), partial (2–4/5), or minimal (0–1/5).

### Graphical Figures

All figures were created using R software (version 4.2.2) and RStudio version 2026.05.0+218.**^(15)^**

### Ethical Considerations

This systematic review relied entirely on data extracted from existing, peer-reviewed publications and did not involve the collection of any new or identifiable human data, hence Institutional review board (IRB) approval was not required.

## Results

### Study Selection and Characteristics

A total of 2048 studies were included initially, of which 422 were duplicates identified by Covidence. Subsequently, 1626 abstracts underwent screening, and 130 were finalized for full text review. After review of these articles, 18 studies were included. Common reasons for exclusion were the absence of a population or health system denominator (n = 104). **Figure 1** shows the PRISMA flowchart for our review.

**Figure 1.**
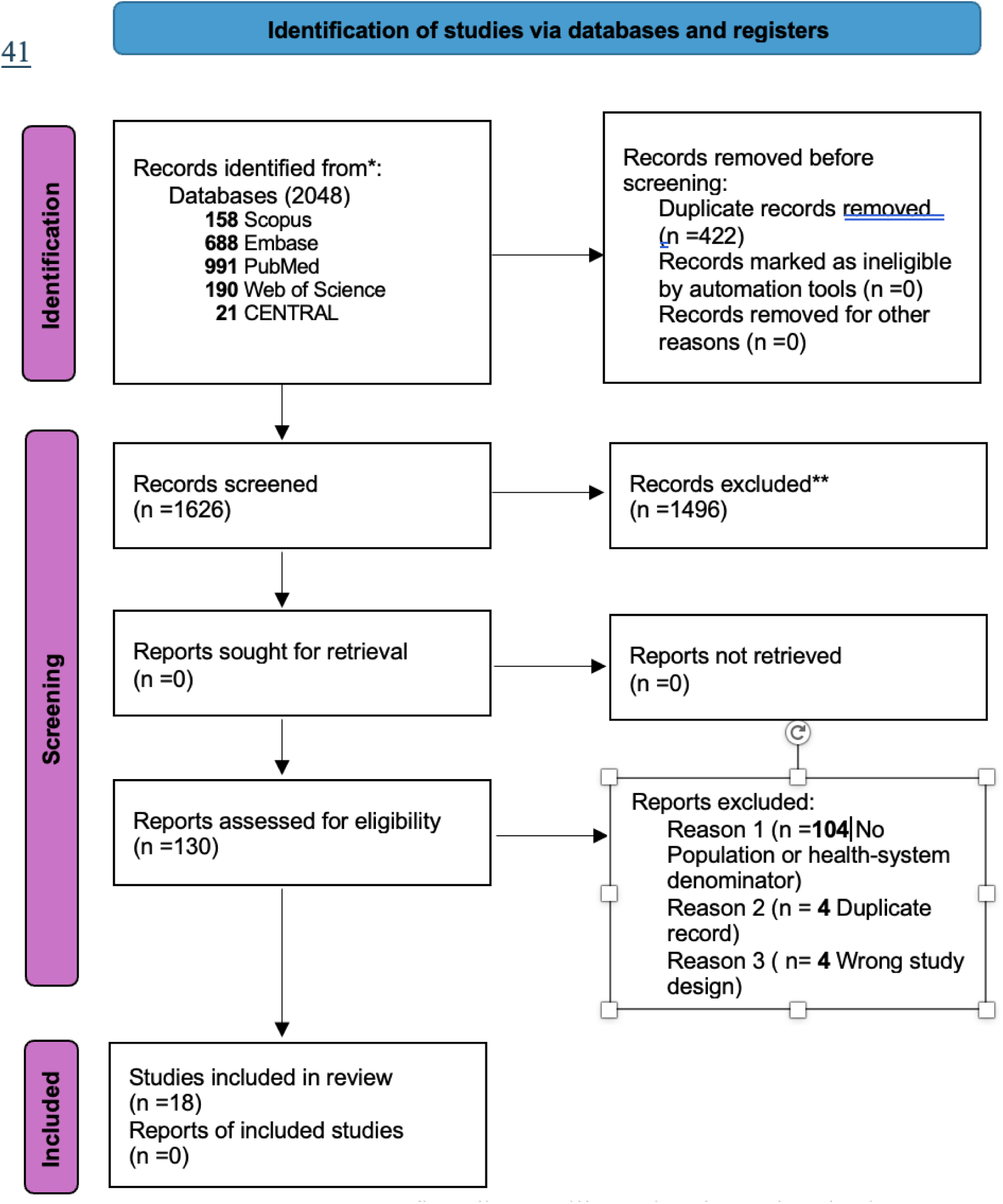
PRISMA flow diagram illustrating the study selection process.

Studies included were conducted between 2013-2026. The reported data spanned from 2000 to 2024, with a median observation window of 9 years (ranging from 4-16 years). These studies were conducted in a wide range of geographical distributions, including South America, North America, and Europe. The most frequent geographical location was the United States. Most of the studies were retrospective observational, with four cross-sectional studies. **(Table 1)**

**Table 1.** Characteristics of the included studies contributing burden or trend estimates.

| Source type | Study | Country | Data source | Design | Period | Classes<sup>a</sup> | n<sup>b</sup> |
| --- | --- | --- | --- | --- | --- | --- | --- |
| **National administrative** | Peery 2018 | USA | HCUP National Inpatient Sample | Repeated cross-sectional | 2000-2014 | N1, N2 | 13 |
|  | Kaltenbach 2025 | USA | VA Surgical Quality Improvement Program | Retrospective cohort | 2000-2015 | N2 | 4 |
|  | Vu 2021 | USA | Medicare, 100% MedPAR | Retrospective cohort | 2010-2015 | N1, N2 | 5 |
|  | Sakowitz 2023 | USA | HCUP National Inpatient Sample | Retrospective cohort | 2012-2019 | N1, N2 | 6 |
| **Surgical quality registry** | Ozgur 2022 | USA | ACS-NSQIP | Retrospective cohort | 2013-2019 | N2 | 10 |
|  | Sharma 2026 | USA | ACS-NSQIP | Retrospective cohort | 2016-2022 | N2 | 6 |
| **National pathology registry** | Bronzwaer 2018 | Netherlands | PALGA Dutch pathology registry | Retrospective multicentre cohort | 2005-2015 | N4 | 12 |
| **National registry** | Nilsen 2024 | Norway | Norwegian Patient Registry | Repeated cross-sectional | 2008-2021 | N1, N3 | 5 |
| **Screening programme registry** | Roy 2016 | France | French population-based screening | Retrospective cohort | 2003-2012 | N3 | 2 |
|  | Lee 2013 | United Kingdom | English Bowel Cancer Screening Programme | Retrospective observational cohort | 2006-2009 | N3, N4 | 4 |
|  | Paszat 2021 | Canada | ColonCancerCheck, Ontario | Population-based retrospective cohort | 2008-2017 | N3, N4 | 6 |
|  | Rodrigues 2020 | France | Departmental screening database SDDC 87 | Population-based retrospective analysis | 2012-2017 | N3, N4 | 6 |
|  | Cubiella 2021 | Spain | Galician CRC screening programme | Retrospective multicentre cohort | 2013-2019 | N3, N4 | 11 |
| **Integrated health system** | Alam 2022 | USA | Kaiser Permanente Northern California | Retrospective cohort | 2008-2018 | N3 | 11 |
| **Federated EHR network** | Alsakarneh 2026 | USA | TriNetX federated EHR network | Retrospective cohort | 2013-2023 | N2 | 11 |
|  | Kilani 2026 | USA | TriNetX federated EHR network | Retrospective cohort | 2014-2024 | N3, N4 | 5 |
| **Single-centre institutional** | Marres 2017 | Netherlands | Institutional database, Amsterdam | Retrospective cohort | 2009-2014 | N2 | 2 |
|  | Kawaguti 2023 | Brazil | Institutional database, Sao Paulo | Single-centre retrospective cohort | 2016-2019 | N2, N3 | 3 |

<sup>a</sup> N1 = population at risk; N2 = all colorectal resections; N3 = all polyps or lesions detected; N4 = all colonoscopies or screening episodes. Estimates are never combined across classes.
<sup>b</sup> Number of separate estimate rows the study contributes; a study reporting two denominators across three periods contributes six.

Data sources were heterogeneous **(Table 1)**, spanning from national registries to an integrated health system database. Four studies were judged to be nationally representative, one partially so, and 12 were not. 1 study did not report enough detail to judge its representativeness.

Two pairs of studies had the same databases and hence were not treated as independent observations. These included i) Peery 2018 and Sakowitz 2023 analyzing the Healthcare Cost and Utilization Project National Inpatient Sample (HCUP-NIS) and ii) Ozgur 2022 and Sharma 2026, both analyzing the American College of Surgeons – National Surgical Quality Improvement (ACS-NSQIP) database (**Appendix 3**). Both reports were retained for descriptive trend comparison because they used different study periods and/or analytic definitions; they were treated as non-independent reports of the same underlying data source and were not counted as independent evidence.

### Denominators used across studies

Of the 18 studies included, 122 separate burden estimates for EUPs were reported, which were assigned to a denominator class determined a priori as described in our methodology **(Table 2).** 21 estimates across four studies were classified as N1, expressing the burden against the total population at risk. 39 estimates were reported across 9 studies for all colorectal resections (N2); 36 estimates from 9 studies were reported for all polyps or lesions detected on colonoscopy (N3), and 26 estimates from 6 studies for all colonoscopy episodes (N4). Ten out of the 18 studies reported estimates for more than one class, and no study reported estimates for all 4 classes.

**Table 2.**
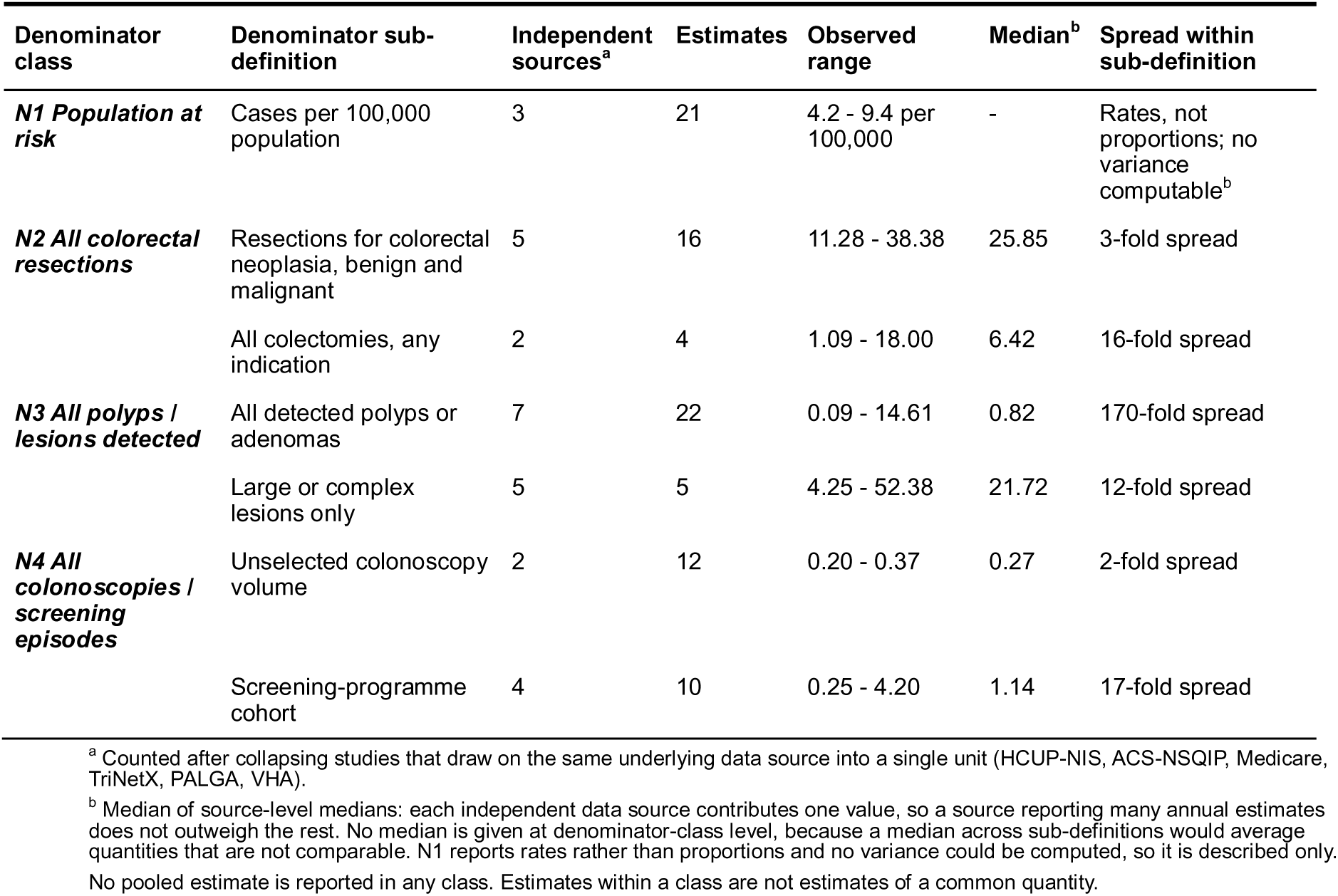
Burden of surgery for endoscopically unresectable colorectal polyps, by denominator class and denominator sub-definition.

| Denominator class | Denominator sub-definition | Independent sources<sup>a</sup> | Estimates | Observed range | Median<sup>b</sup> | Spread within sub-definition |
| --- | --- | --- | --- | --- | --- | --- |
| ***N1 Population at risk*** | Cases per 100,000 population | 3 | 21 | 4.2 - 9.4 per 100,000 | - | Rates, not proportions; no variance computable<sup>b</sup> |
| ***N2 All colorectal resections*** | Resections for colorectal neoplasia, benign and malignant | 5 | 16 | 11.28 - 38.38 | 25.85 | 3-fold spread |
|  | All colectomies, any indication | 2 | 4 | 1.09 - 18.00 | 6.42 | 16-fold spread |
| ***N3 All polyps / lesions detected*** | All detected polyps or adenomas | 7 | 22 | 0.09 - 14.61 | 0.82 | 170-fold spread |
|  | Large or complex lesions only | 5 | 5 | 4.25 - 52.38 | 21.72 | 12-fold spread |
| ***N4 All colonoscopies / screening episodes*** | Unselected colonoscopy volume | 2 | 12 | 0.20 - 0.37 | 0.27 | 2-fold spread |
|  | Screening-programme cohort | 4 | 10 | 0.25 - 4.20 | 1.14 | 17-fold spread |

<sup>a</sup> Counted after collapsing studies that draw on the same underlying data source into a single unit (HCUP-NIS, ACS-NSQIP, Medicare, TriNetX, PALGA, VHA).
<sup>b</sup> Median of source-level medians: each independent data source contributes one value, so a source reporting many annual estimates does not outweigh the rest. No median is given at denominator-class level, because a median across sub-definitions would average quantities that are not comparable. N1 reports rates rather than proportions and no variance could be computed, so it is described only.
No pooled estimate is reported in any class. Estimates within a class are not estimates of a common quantity.

However, notably, assignment to the same class did not allow for two estimates to be statistically comparable. This is because 28 distinct denominator descriptions were reported within N2, with 21 in N3, 14 in N1, and 12 in N4. **(Appendix 3)**. As an example, within N2, the denominator varies from all colectomies performed for any indication to all elective partial colectomies for colonic neoplasm. Therefore, all narrative synthesis is stratified by class, and within classes, by denominator definition.

### Burden within each denominator class

Pooled estimates are not reported for any denominator class as a whole, because heterogeneity in denominator definition meant that estimates within a class were not of a common quantity; pooling was therefore restricted to denominator sub-definitions and is reported separately below. Every proportion was recomputed from the published numerator and denominator before analysis. In total, 69 of 122 estimates provided the numerator and denominator pair required for comparison. The remaining 53 were described qualitatively, because they reported no numeric estimate, were expressed as population rates, lacked a numerator or denominator, or duplicated another estimate of the same patients **(Appendix 3).** Therefore, summary values were reported as observed ranges and as medians within each denominator sub-definition. **(Table 2**, **Figure 2)**

**Figure 2.**
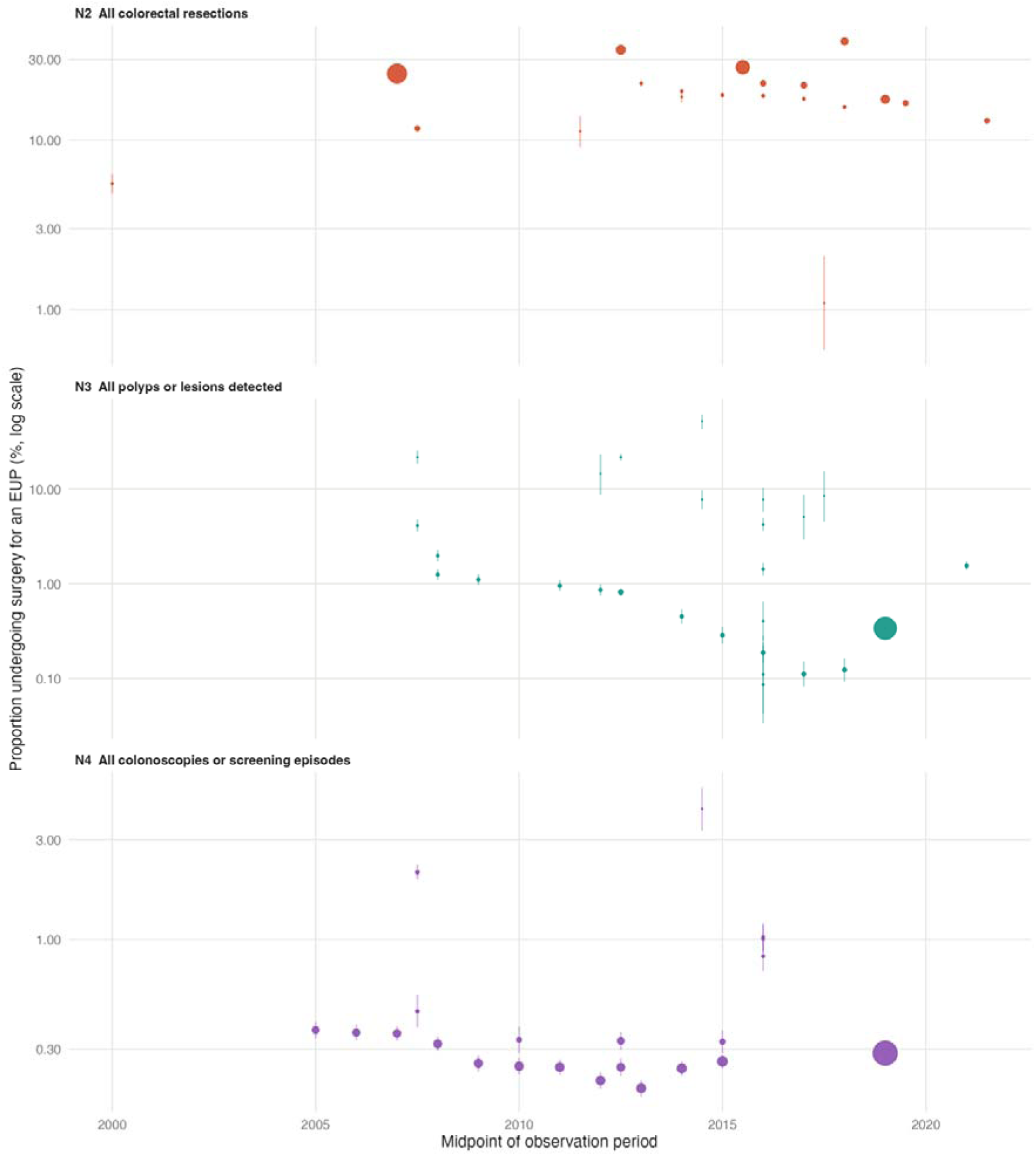
Reported burden estimates by denominator class and observation period. Each point is one of the 69 estimates for which both a numerator and a denominator were reported, plotted against the midpoint of its observation period: 20 estimates against all colorectal resections (N2), 27 against all polyps or lesions detected (N3), and 22 against all colonoscopies or screening episodes (N4). Vertical bars are binomial 95% intervals (descriptive) calculated from the reported numerator and denominator. Point area is proportional to the size of the denominator. The vertical axis is scaled independently in each panel. Estimates expressed as population rates (N1) are not shown here, as no variance could be computed from the reported figures.

**Figure 3.**
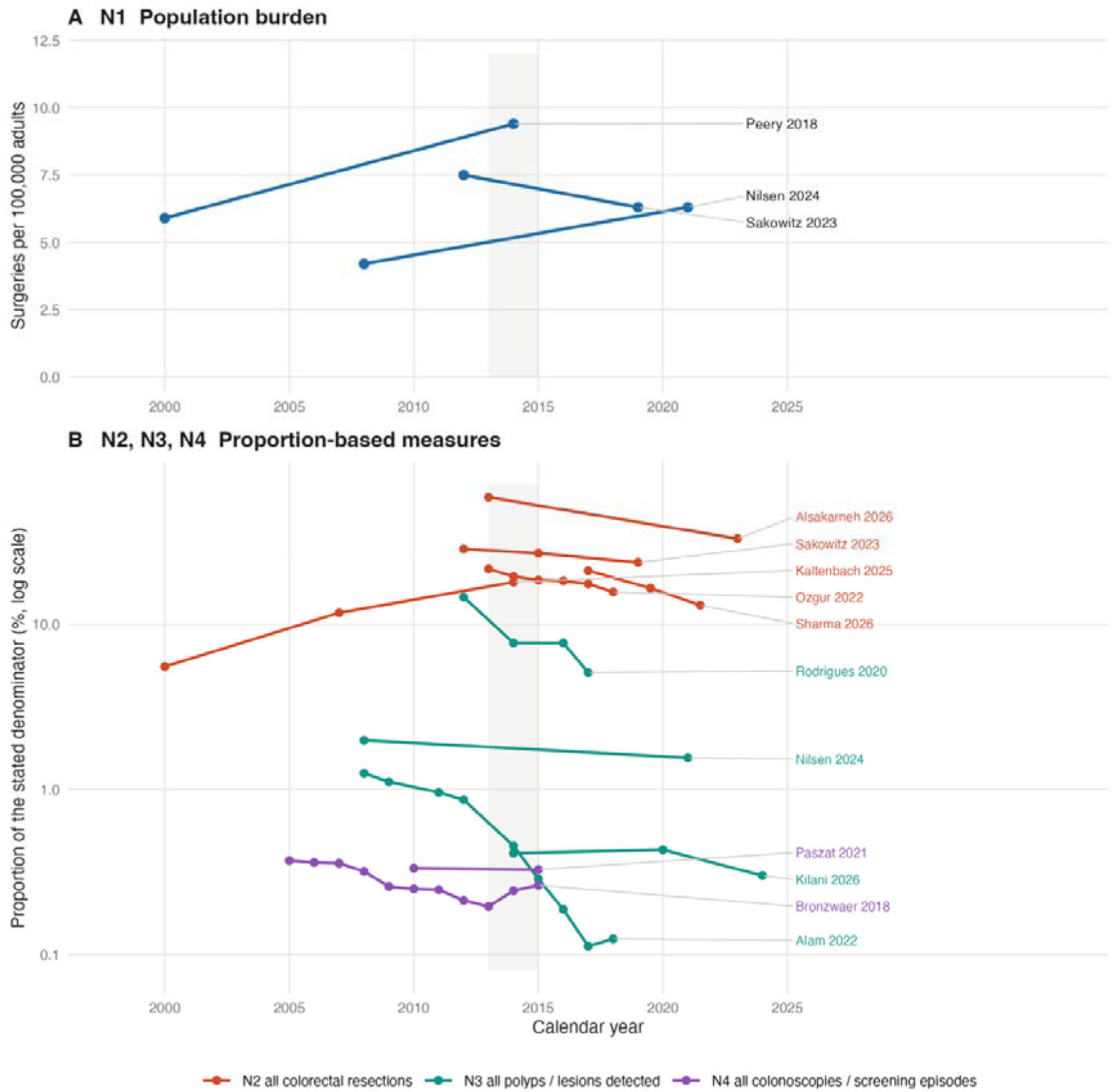
Temporal trends by study and denominator class. Fourteen series from 12 studies are shown; a series is drawn only where a single study reported the same measure at two or more time points. The shaded band marks 2013–2015, around which the majority of series change direction. These studies were not pooled together because of denominator heterogeneity.

Total population at risk (N1): Four studies using 3 independent data sources reported the burden as a rate per 100,000 population. These ranged from 4.2 to 9.4 per 100,000 adults. No study reported a numerator and denominator pair that could be used to compute a variance, and hence only a narrative description was performed.

All colorectal resections (N2): Twenty proportions from seven different sources ranged from 1.09% to 38.38%. This spread was also stratified by denominator sub-definition. Where the denominator comprised resections for colorectal neoplasia alone, sixteen estimates from five sources ranged from 11.28% to 38.38%, with a median across sources of 25.85%. Where it comprised all colectomies performed for any indication, four estimates from two sources ranged from 1.09% to 18.00%, with a median of 6.42%. The lowest estimate within this class (1.09%, Kawaguti 2023) came from an established single-center Endoscopic Submucosal Dissection (ESD) service, whereas the highest came from an electronic healthcare network. (38.38%, Alsakarneh 2026)

All polyps or lesions detected (N3): 27 different proportions were found from nine different data sources, ranging from 0.086% to 52.38%. This was the widest spread observed in this review. This wide range was attributed to restriction of the denominators by authors using lesion size or complexity. Where the denominator comprised all detected polyps or adenomas, twenty estimates from seven sources ranged from 0.09% to 14.61%, with a median across sources of 0.82%. Where it was restricted to large or complex lesions, five estimates from five sources ranged from 4.25% to 52.38%, with a median of 21.72%

All colonoscopies or screening events (N4): 22 analyzable estimates from six independent data sources, ranging from 0.20% to 4.2%, the most consistent range observed in this review. Where the denominator comprised unselected colonoscopy volume, 12 estimates from two national datasets (One European pathology registry and a North American federated network) ranged from 0.20% to 0.37%, with a median of 0.27%. When it comprised a screening programme cohort defined by a positive fecal blood test (10 estimates from 4 sources), estimates ranged from 0.25% to 4.20% (median 1.14%).

### Pooled estimates within denominator sub-definitions

Three of the six denominator sub-definitions contained estimates from at least three independent data sources and were pooled **(Table 5**, **Figure 5)**. Among resections for colorectal neoplasia (N1), five sources gave a pooled proportion of 23.79% (95% CI 12.27 to 41.06), with a 95% prediction interval of 3.22% to 74.55% and a between-study variance of 0.411. Among all detected polyps or adenomas (N2), seven sources gave a proportion of 1.15% (95% CI 0.31 to 4.21), prediction interval 0.02% to 37.19%, between-study variance 2.040. Among large or complex lesions only (N3), five sources gave a proportion of 17.11% (95% CI 4.36 to 48.30), prediction interval 0.31% to 93.22%, between-study variance 1.445. Among screening-programme cohorts (N4), four sources gave a proportion of 0.84% (95% CI 0.12 to 5.80), prediction interval 0.00% to 76.98%, between-study variance 1.542.

Two sub-definitions contained only two independent data sources and were not pooled: all colectomies performed for any indication (1.09% and 11.74%) and unselected colonoscopy volume (0.262% and 0.287%).

I-squared exceeded 99% in every pooled sub-definition. With denominators of this size the within-study variance is negligible and I-squared approaches a maximum by construction, so the prediction intervals are the informative measure. In all four pooled sub-definitions the prediction interval spanned more than an order of magnitude, and in three of the four more than two orders of magnitude, indicating that a new study drawn from this literature could plausibly report almost any value. The narrowest interval was obtained for resections for colorectal neoplasia, the sub-definition with the most tightly specified denominator, and even there it spanned approximately 23-fold.

Restricting estimates to a single sub-definition reduced between-study variance most where sub-definitions differed most in the lesions they admitted. Among polyp-based denominators the between-study variance fell from 3.823 for the class as a whole to a source-weighted 1.792 within sub-definitions, a reduction of 53%. The corresponding reductions were 32% for resection-based and 18% for colonoscopy-based denominators.

### Sources of disagreement between estimates

Variation between reported estimates appeared to reflect differences in denominator definition, study setting, and EUP ascertainment. These discrepancies were also observed within individual studies. Seven studies reported the same numerator against two or more denominators (**Table 3**, **Figure 4**). As an example, Paszat 2021 reported a count of 420 surgical cases for EUPs against four denominators, yielding estimates of 0.25%, 0.33%, 0.82% and 21.73%, which is a wide range. Similarly, Lee 2013 reported 121 cases against two denominators, resulting in estimates of 0.46% and 21.72%. In such instances, the surgical events were completely identical, but the denominators used made the estimate variable.

**Table 3.** Estimates generated from a single numerator within one study, by the denominator applied.

| Study and numerator | Denominator (n) | Denominator sub-definition | Class | Estimate (%) | Largest / smallest |
| --- | --- | --- | --- | --- | --- |
| **Paszat 2021 (numerator = 420)** | 1,933 | Large or complex lesions only | N3 | 21.73 | 88.3-fold |
|  | 51,310 | All detected polyps or adenomas | N3 | 0.82 |  |
|  | 127,872 | Screening-programme cohort | N4 | 0.33 |  |
|  | 170,670 | Screening-programme cohort | N4 | 0.25 |  |
| **Lee 2013 (numerator = 121)** | 557 | Large or complex lesions only | N3 | 21.72 | 47.7-fold |
|  | 26,552 | Screening-programme cohort | N4 | 0.46 |  |
| **Kawaguti 2023 (numerator = 9)** | 106 | Large or complex lesions only | N3 | 8.49 | 7.8-fold |
|  | 825 | All colectomies, any indication | N2 | 1.09 |  |
| **Rodrigues 2020 (numerator = 66)** | 856 | All detected polyps or adenomas | N3 | 7.71 | 1.8-fold |
|  | 1,571 | Screening-programme cohort | N4 | 4.20 |  |
| **Cubiella 2021 (numerator = 158)** | 11,053 | All detected polyps or adenomas | N3 | 1.43 | 1.4-fold |
|  | 15,707 | Screening-programme cohort | N4 | 1.01 |  |
| **Cubiella 2021 (numerator = 4)** | 3,630 | All detected polyps or adenomas | N3 | 0.11 | 1.3-fold |
|  | 4,654 | All detected polyps or adenomas | N3 | 0.09 |  |
| **Kilani 2026 (numerator = 5,750)** | 1,693,869 | All detected polyps or adenomas | N3 | 0.34 | 1.2-fold |
|  | 2,003,307 | Unselected colonoscopy volume | N4 | 0.29 |  |

**Figure 4.**
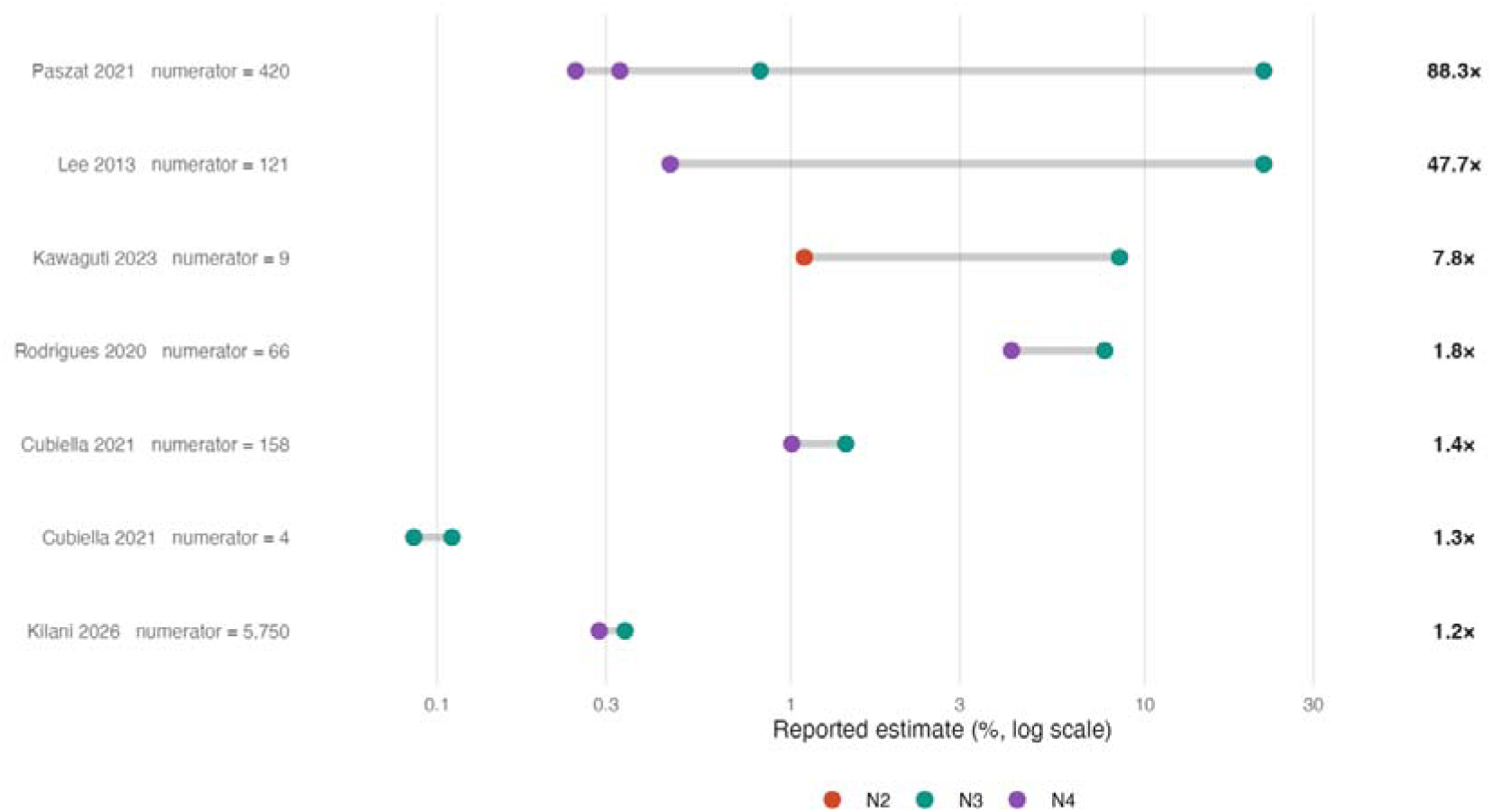
Divergence of estimates generated from a single numerator. Sixteen estimates from six studies are shown, arranged as seven rows. Within any row the surgical events counted are identical; the entire divergence is a consequence of denominator choice within the same row. Paszat 2021 reported a single count of 420 surgical cases against four denominators, yielding estimates from 0.25% to 21.73%.

**Figure 5.**
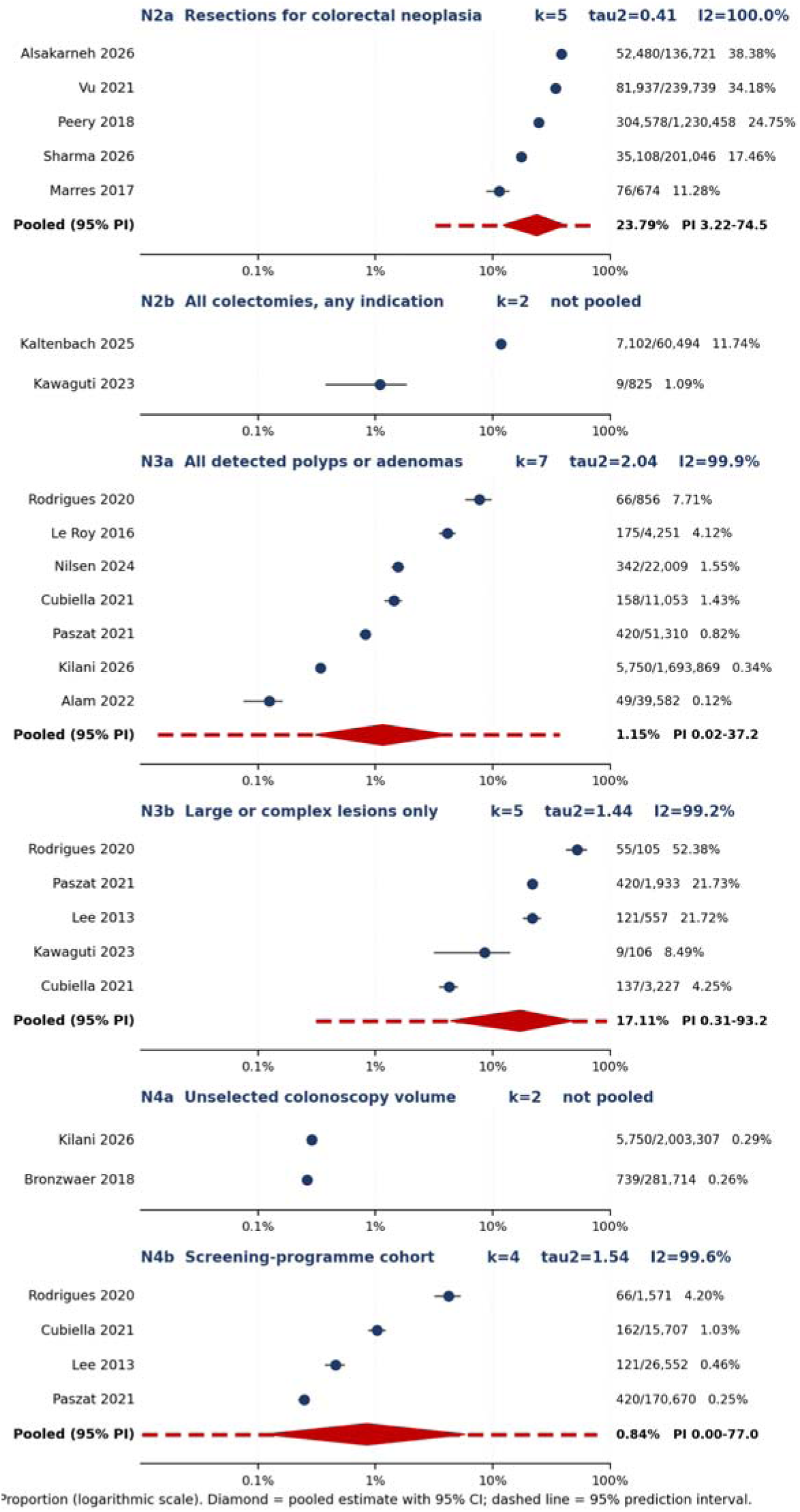
Pooled proportions with 95% prediction intervals, by denominator sub-definition. Random-effects models on the logit scale, one estimate per independent data source after collapsing studies that share an underlying dataset. Circles are individual source estimates with binomial 95% intervals; the red diamond is the pooled proportion with its Hartung-Knapp 95% confidence interval; the dashed red line is the 95% prediction interval. The horizontal axis is logarithmic. N2b and N4a are shown without a pooled estimate because each contains only two independent sources. In every pooled sub-definition the prediction interval spans more than an order of magnitude, indicating that a future study drawn from this literature could report almost any value.

This same disagreement was observed within classes. Within N4, the twelve reported values extracted from all colonoscopy volumes spanned only 0.195% to 0.37%, across two independent national datasets on different continents, whereas the class as a whole spanned 0.20% to 4.20%. When the denominators were identical, the reported estimates were closely comparable.

### Completeness of unresectability designation

Reporting of criteria used to determine an EUP to be labelled unresectable against five items decided a priori was completed for 17 of the 18 studies. 1 study did not provide enough information to be ascertained against these criteria. **(Appendix 3)**

All 17 studies reported whether ascertainment of resectability was applied retrospectively or prospectively. Seven stated criteria for unresectability at all, and seven studies stated the point in the referral pathway where the lesion was labelled to be unresectable. Five studies stated the criteria with sufficient operational clarity to be applied independently, and five studies stated the individual designating the lesion to be an EUP. Summing it up, only one study scored 5/5 (complete), nine scored 2-4/5 (partial), and seven scored 0-1/5 (minimal).

Seven of the 17 scored studies stated any criteria for endoscopic unresectability. The remaining ten reported a rate of endoscopically unresectable polyps without defining EUPs. Studies with minimal ascertainment reporting were consistently administrative or registry-based, and reported the highest estimates within the N2 class. Moderator analysis of ascertainment could not be conducted because only one study achieved a complete score. Moreover, another reason analysis could not be conducted was because of the collinearity between data source and ascertainment completeness.

### Temporal trends

16 trend estimates were reported across all four denominator classes, as is seen in **Table 4 and Figure 3**. Trends reported by studies were displayed alongside one another and were not pooled together.

**Table 4.**
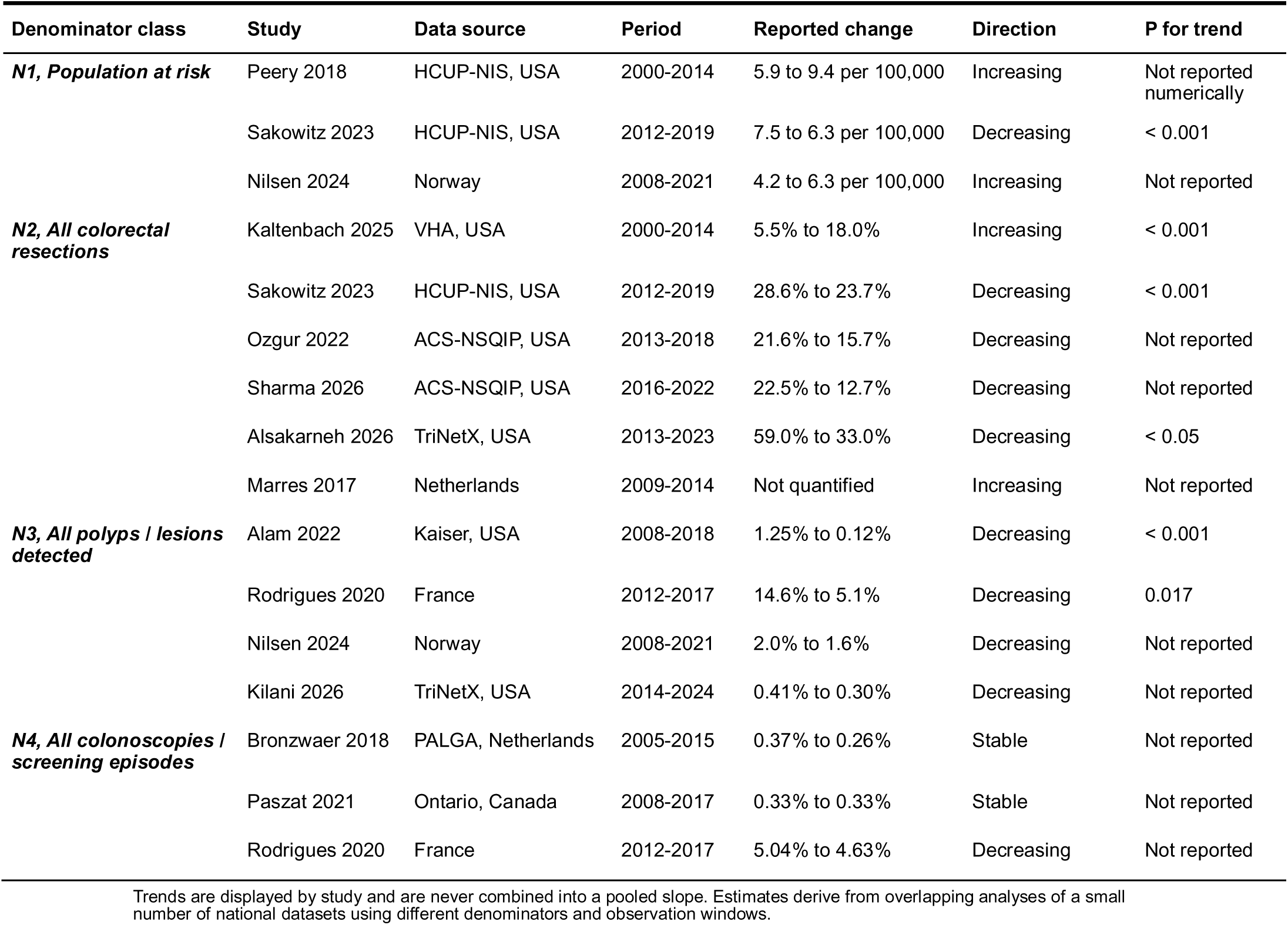
Reported temporal trends, by study and denominator class.

**Table 5.** Pooled proportions within denominator sub-definitions.

| Denominator sub-definition | Independent sources (k) | Observed range (%) | Pooled % (95% CI) | 95% prediction interval (%) | Between-study variance | I<sup>2</sup> (%) |
| --- | --- | --- | --- | --- | --- | --- |
| N2a Resections for colorectal neoplasia | 5 | 11.28 to 38.38 | 23.79 (12.27 to 41.06) | 3.22 to 74.55 | 0.411 | 100.0 |
| N2b All colectomies, any indication | 2 | 1.09 to 11.74 | Not pooled (k<3) | - | - | - |
| N3a All detected polyps or adenomas | 7 | 0.12 to 7.71 | 1.15 (0.31 to 4.21) | 0.02 to 37.19 | 2.040 | 99.9 |
| N3b Large or complex lesions only | 5 | 4.25 to 52.38 | 17.11 (4.36 to 48.30) | 0.31 to 93.22 | 1.445 | 99.2 |
| N4a Unselected colonoscopy volume | 2 | 0.26 to 0.29 | Not pooled (k<3) | - | - | - |
| N4b Screening-programme cohort | 4 | 0.25 to 4.20 | 0.84 (0.12 to 5.80) | 0.00 to 76.98 | 1.542 | 99.6 |

Reported directions over time were not consistent, and the inconsistency observed was secondary to the time window rather than the denominator used or the setting. Studies that reported estimates before 2014 reported increases in surgical intervention for EUPs; Kaltenbach 2025 reported a rise from 5.5% to 18.0% of all colectomies between 2000 and 2014, and Peery 2018 a rise from 5.9 to 9.4 per 100,000 adults over the same period. On the other hand, studies that reported estimates from 2012 to 2024 reported decreases, for every denominator class reported. These included Sakowitz 2023 (28.6% to 23.7%), Ozgur 2022 (21.6% to 15.7%), Sharma 2026 (22.5% to 12.7%), Alsakarneh 2026 (59.0% to 33.0%), Alam 2022 (1.25% to 0.12%), Rodrigues 2020 (14.6% to 5.1%) and Kilani 2026 (0.41% to 0.30%).

Two studies reported stable estimates over time: Bronzwaer 2018, which observed all colonoscopy events (N4) from 2005 to 2015. The other one was Paszat 2021, across 2008 to 2017, also observing colonoscopy events (N4).

Moreover, it was also observed that within a single population, trends moved in different trajectories. Nilsen 2024 reported a rise in the population rate from 4.2 to 6.3 per 100,000 between 2008 and 2021, while the proportion of EUPs managed surgically fell from 2.0% to 1.6% over the same period.

### Risk of Bias Assessment

Risk of bias was assessed for 17 of the 18 studies using the JBI Critical Appraisal Checklist **(Appendix 4)**. Roy 2016 was not assessed because of insufficient data for a judgement to be made. The final verdict included two studies at low risk of bias, six at moderate risk, and nine at high overall risk of bias. All studies described their subjects and study settings. 14 studies had an adequately large sample size, owing to the database and registry data. Valid methods of ascertainment were judged adequately in eight studies, and the condition was measured in a standard way in only three studies.

Of the five supplementary metrics used for database/registry studies, the sampling frame was representative in six studies. Procedural and diagnostic codes were used in full in eight of the 12 studies using code-based ascertainment. The denominator was stable in four and unstable in eight studies, primarily because it was evaluated by colonoscopy volume, so a reported proportion could be altered without the number of surgeries being affected. A coding revision falling within the observation window was fully addressed in three studies.

### Certainty of Evidence

Certainty of evidence was evaluated using the GRADE framework (adapted for questions of event rate), separately for each denominator class **(Appendix 4)**. All four classes were downgraded for risk of bias and for indirectness. Inconsistency was rated very serious for N1, N2 and N3, and serious for N4. Imprecision was rated not serious across all four classes, as individual estimates in this study were precisely determined. Certainty was therefore very low for the population at risk (N1), for all colorectal resections (N2) and for all detected polyps or lesions (N3), and low for all colonoscopies or screening episodes (N4).

## Discussion

This systematic review included 18 studies reporting the burden of surgery for endoscopically unresectable colorectal polyps against a population or health-system denominator, contributing to 122 separate estimates. Estimates of the same nominal quantity varied widely, ranging from 1.09% to 38.38% of colorectal lesions and 0.09% to 52.38% of polyps detected. This dispersion of data could not be explained by country, calendar period, or the type of health system, but rather by how each study constructed its denominator and its numerator, and more interestingly, by how they defined the case. 75 distinct denominator definitions were in use across the four classes, with 28 of them within the single class “all colorectal resections”. Where denominators matched, estimates agreed closely. Furthermore, only 1 of 17 studies fully reported the criteria used to designate a polyp as endoscopically unresectable, with an additional 7 studies minimally reporting the same criteria. A summary estimate was not defensible across different denominator classes and hence is not included in this review.

The harm of surgery for benign unresectable polyps is well established. de Neree tot Babberich et al. pooled 26 studies with a combined 139,897 patients and found a 30-day adverse event rate of 24% and a mortality rate of 0.7%**^(16)^**. This study highlighted the morbidity associated with surgery but did not answer how often the operation is performed, nor against what denominator. The same study acknowledged but did not measure the fact that the criteria for endoscopic unrespectability varied greatly between health systems. Our study takes this observation as its object. Its contributions are a taxonomy of the denominators in use, and a quantified assessment of how completely the case definition is reported in this literature.

Four different denominator classes are in circulation, each answering a different question. The population rate is not affected by surgical volume; the proportion of colorectal resections is sensitive to changes in operating for other indications; the proportion of detected polyps measures referral behavior after detection; and the proportion of colonoscopies is sensitive to endoscopic volume, which rose substantially across every study period examined. Proof that these denominators cannot be interchanged can be represented within a single population; Nilsen et al reported a rise in population rate from 4.2 to 6.3 per 100,000 between 2008 and 2021 according to the Norwegian national registry. However, the proportion of detected polyps managed surgically fell from 2.0% to 1.6% within the same time span**^(17)^**. Heterogeneity of the data was not confined to the choice between classes. Within “all colorectal resections”, estimates restricted to resections for colorectal neoplasia ranged from 11.28% to 38.38%, with a median value of 25.85%. Whereas those using all colectomies for any indication ranged from 1.09% to 18.00%, with a median value of just 6.42%. This is a difference in denominator only, and not one of practice.

The strongest evidence that the denominator drives the disagreement comes from within individual studies themselves. 7 of the 18 included studies reported a single numerator against two or more denominators. Paszat et al reported 420 surgical cases as 0.25%, 0.33%, 0.82%, and 21.73%, an 88.3-fold spread generated by the same 420 cases**^(18)^**. Similarly, Lee et al reported 121 cases as 0.46% and 21.72%, a 47.7-fold spread**^(19)^**. The converse relation also holds. Twelve estimates of surgery per unselected colonoscopy, drawn from two independent national datasets on different continents, spanned only 0.20% to 0.37%**^(10,20)^**. When the denominator is held constant, estimates from unrelated health systems converge to within a factor of two. This is the central finding of the review, and it means that cross-study comparison of this burden is uninterpretable unless the denominator definition is reproduced alongside the estimate.

The case definition compounds this problem. The criteria for endoscopic unresectability were stated in only 7 of 17 assessable studies; the remaining ten only reported a rate of surgery for endoscopically unresectable polyps without defining the term itself. Five studies state their criteria with sufficient operational precision for independent application, and five identified who made the designation for the polyp to be deemed unresectable. One study satisfied all five of the reporting items, nine satisfied two to four, and seven satisfied one or none of them. Every study classified in the minimal category was an analysis of administrative or registry data, and these studies reported the highest estimates within the colorectal-resection class, consistent with a case definition that captured any benign neoplasm undergoing resection rather than a lesion judged unresectable by a clinician. The relationship between cohort size and definitional precision is therefore inverse: the studies large enough to estimate a population burden are least able to actually define the construct that they are counting. We were unable to formally test ascertainment completeness as a moderator, since only one study achieved a complete score and because the data source and ascertainment completeness were collinear.

By contrast, once the observation window was considered, the trend direction proved to be coherent. Sixteen trend estimates were reported, and their apparent disagreement tracked the period observed rather than the denominator or the setting. Studies whose data extended before 2014 reported increases from 5.5% to 18.0% of colectomies between 2000 and 2014**^(21)^**, and from 5.9 to 9.4 per 100,000 adults over the same period**^(22)^**. However, studies spanning 2012 to 2024 reported decreases in every denominator class, across seven independent data sources in four countries and on both population and proportional denominators**^(9,10,23–27)^**. Two national screening datasets reported stability across windows straddling that point**^(18,20)^**. Together, these findings suggest that there was a genuine inflection in practice around the 2012–2014-timeframe, after which surgery for these lesions has declined noticeably. Nonetheless, this should be interpreted cautiously, as the trend estimates derive from overlapping analysis of a small number of national datasets, with most reporting no trend statistic, and in two of them the population denominator was held constant across the series, so that the reported incidence trend is a numerator trend with a fixed divisor.

This review has several limitations that need acknowledgement. Firstly, publication bias could not be assessed, because no denominator class contained ten or more independent data sources once studies that shared an underlying dataset were counted once. Secondly, several sources are not entirely independent: two pairs of included studies analyze the same databases**^(22–25)^**, and these were counted once. Additionally, 53 of the 122 estimates could not enter any quantitative comparison because they did not report any numerator and denominator pair. One study was available only as a published abstract and could not be assessed for reporting completeness. Furthermore, certainty of evidence was very low for three of the four denominator classes. These were rated down principally for inconsistency and indirectness rather than containing imprecise data; the individual estimates are precise, but they are of different quantities. Lastly, the instrument used to assess reporting completeness has not been externally validated as it has been developed for this review specifically.

With regard to future directions, four implications should follow. For practice, the current European guidance recommends en bloc techniques where superficial invasive carcinoma is suspected and identifies lesion size above 40mm as a basis for referral to an expert interventional center. Factors such as ileocaecal valve or dentate line involvement, previous failed resection, and a high difficulty score based on the size, morphology, site, and access of the polyp are all bases for directing a patient to a high-volume interventional endoscopy center, yet almost none of the studies included in this review document the application of any such criterion before surgical referral**^(28)^**. For the design of audits and registries, administrative coding should not be relied on to express a resectability judgement. The health systems that wish to monitor this quantity should be capturing the decision rather than the operation. A dataset, at minimum, should record the lesion size and morphology, whether or not endoscopic resection was attempted, and if expert opinion was sought before the patient was referred for surgery. Additionally, the residual population of lesions that remain unsuitable for resection represents a potential target for robotic endoluminal platforms. Factors leading to referral for surgery, including large, sessile, or anatomically challenging polyps, are exactly those challenges which robotic platforms could resolve. Future research should explore the possibility of developing robotic endoscopic platforms capable of deeper margin resections and precise tissue manipulation, as they could reduce the need for surgical removal of endoscopically unresectable polyps and potentially expand organ-preserving treatment**^(29)^**. For future research, any study reporting this burden should present the numerator and denominator as counts with an explicit definition of each of them and state whether lesions operated after a failed endoscopic attempt are included and report the criteria by which unresectability was determined and by whom. Without such reporting, future studies will continue to simply add volume to this literature without contributing additional knowledge.

## Author contributions

MMA contributed to conceptualization, methodology, software, formal analysis, investigation, data curation, writing - original draft, writing - review and editing, and project administration. MTB contributed to methodology, software, formal analysis, investigation, data curation, writing - original draft, writing - review and editing. MS contributed to methodology, writing - review and editing, visualization, validation, and project administration. AKu, NK, AC contributed to methodology, validation, visualization, and writing - review and editing. AKh contributed to conceptualization, methodology, writing - review and editing, and supervision.

## Data Availability

All data produced are available online at PubMed, Scopus, Web of Science, Cochrane CENTRAL and Embase. Our search strings are in the appendices.

## Acknowledgements

None

## References

1. Almadi MajidA, Alharbi O, Azzam N, Wadera J, Sadaf N, Aljebreen Abdulrahman M. Prevalence and characteristics of colonic polyps and adenomas in 2654 colonoscopies in Saudi Arabia. Saudi J Gastroenterol. 2014;20(3):154. doi:10.4103/1319-3767.132986

2. Wong MCS, Huang J, Huang JLW, Pang TWY, Choi P, Wang J, et al. Global Prevalence of Colorectal Neoplasia: A Systematic Review and Meta-Analysis. Clin Gastroenterol Hepatol. 2020 Mar;18(3):553–561.e10. doi:10.1016/j.cgh.2019.07.016

3. Cannon-Albright LA, Bishop DT, Samowitz W, DiSario JA, Lee R, Burt RW. Colonic polyps in an unselected population: prevalence, characteristics, and associations. Am J Gastroenterol. 1994 Jun;89(6):827–31. PubMed PMID: 8198089.

4. Rutter MD, Chattree A, Barbour JA, Thomas-Gibson S, Bhandari P, Saunders BP, et al. British Society of Gastroenterology/Association of Coloproctologists of Great Britain and Ireland guidelines for the management of large non-pedunculated colorectal polyps. Gut. 2015 Dec;64(12):1847–73. doi:10.1136/gutjnl-2015-309576

5. Pattarajierapan S, Takamaru H, Khomvilai S. Difficult colorectal polypectomy: Technical tips and recent advances. World J Gastroenterol. 2023 May 7;29(17):2600–15. doi:10.3748/wjg.v29.i17.2600 PubMed PMID: 37213398; PubMed Central PMCID: PMC10198056.

6. Bertelson NL, Kalkbrenner KA, Merchea A, Dozois EJ, Landmann RG, De Petris G, et al. Colectomy for endoscopically unresectable polyps: how often is it cancer? Dis Colon Rectum. 2012 Nov;55(11):1111–6. doi:10.1097/DCR.0b013e3182695115 PubMed PMID: 23044670.

7. Patel M, Haque M, Kohli D, Mutha P, Shah SA, Fernandez L, et al. Endoscopic resection reduces morbidity when compared to surgery in veterans with large and complex colorectal polyps. Surg Endosc. 2021 Mar;35(3):1164–70. doi:10.1007/s00464-020-07482-y PubMed PMID: 32166551.

8. Wickham CJ, Wang J, Mirza KL, Noren ER, Shin J, Lee SW, et al. “Unresectable” polyp management utilizing advanced endoscopic techniques results in high rate of colon preservation. Surg Endosc. 2022 Mar;36(3):2121–8. doi:10.1007/s00464-021-08499-7 PubMed PMID: 33890178.

9. Alsakarneh S, Karna R, Shaukat A, Bilal M. Rates of colorectal surgery in patients with non-malignant colorectal polyps: Results from a nationwide study. Endosc Int Open. 2026;14:a27957563. doi:10.1055/a-2795-7563 PubMed PMID: 41704857; PubMed Central PMCID: PMC12908939.

10. Kilani Y, Madi MY, Mosquera DAG, Bazarbashi AN, McCarty TR, Shah R. Trends in Surgical and Endoscopic Resection Intervention for Non-malignant Colorectal Polyps Over the Last Decade: A Nationwide Analysis. Dig Dis Sci. 2026 Feb;71(2):605–14. doi:10.1007/s10620-025-09372-6 PubMed PMID: 40884699.

11. Page MJ, McKenzie JE, Bossuyt PM, Boutron I, Hoffmann TC, Mulrow CD, et al. The PRISMA 2020 statement: an updated guideline for reporting systematic reviews. BMJ. 2021 Mar 29;n71. doi:10.1136/bmj.n71

12. Covidence systematic review software, Veritas Health Innovation, Melbourne, Australia. Available at www.covidence.org.

13. Munn Z, Moola S, Lisy K, Riitano D, Tufanaru C. Methodological guidance for systematic reviews of observational epidemiological studies reporting prevalence and cumulative incidence data. Int J Evid Based Healthc. 2015 Sep;13(3):147–53. doi:10.1097/XEB.0000000000000054 PubMed PMID: 26317388.

14. Iorio A, Spencer FA, Falavigna M, Alba C, Lang E, Burnand B, et al. Use of GRADE for assessment of evidence about prognosis: rating confidence in estimates of event rates in broad categories of patients. BMJ. 2015 Mar 16;350:h870. doi:10.1136/bmj.h870 PubMed PMID: 25775931.

15. Posit team (2025). RStudio: Integrated Development Environment for R. Posit Software, PBC, Boston, MA. URL http://www.posit.co/.

16. de Neree Tot Babberich MPM, Bronzwaer MES, Andriessen JO, Bastiaansen BAJ, Mostafavi N, Bemelman WA, et al. Outcomes of surgical resections for benign colon polyps: a systematic review. Endoscopy. 2019 Oct;51(10):961–72. doi:10.1055/a-0962-9780 PubMed PMID: 31330557.

17. Nilsen JA, Bernklev L, Bretthauer M, Kalager M, Jodal HC, Løberg M, et al. Surgical treatment of benign colorectal polyps 2008–21. Tidsskr Den Nor Legeforening. 2024 Sep 4;144(10). doi:10.4045/tidsskr.23.0722

18. Paszat LF, Sutradhar R, Luo J, Baxter NN, Tinmouth J, Rabeneck L. Morbidity and mortality after major large bowel resection of non-malignant polyp among participants in a population-based screening program. J Med Screen. 2021 Sep;28(3):261–7. doi:10.1177/0969141320967960 PubMed PMID: 33153368; PubMed Central PMCID: PMC8366188.

19. Lee TJW, Rees CJ, Nickerson C, Stebbing J, Abercrombie JF, McNally RJQ, et al. Management of complex colonic polyps in the English Bowel Cancer Screening Programme. Br J Surg. 2013 Nov;100(12):1633–9. doi:10.1002/bjs.9282 PubMed PMID: 24264787.

20. Bronzwaer MES, Koens L, Bemelman WA, Dekker E, Fockens P, COPOS study group. Volume of surgery for benign colorectal polyps in the last 11 years. Gastrointest Endosc. 2018 Feb;87(2):552–561.e1. doi:10.1016/j.gie.2017.10.032 PubMed PMID: 29108978.

21. Kaltenbach T, Martin L, Yu J, Brooke B, Peche W, Soetikno R, et al. Elective colectomy for treatment of benign colon polyps: National surgical trends, outcomes, and cost analysis. Endosc Int Open. 2025;13:a26895839. doi:10.1055/a-2689-5839 PubMed PMID: 41142263; PubMed Central PMCID: PMC12550741.

22. Peery AF, Cools KS, Strassle PD, McGill SK, Crockett SD, Barker A, et al. Increasing Rates of Surgery for Patients With Nonmalignant Colorectal Polyps in the United States. Gastroenterology. 2018 Apr;154(5):1352–1360.e3. doi:10.1053/j.gastro.2018.01.003 PubMed PMID: 29317277; PubMed Central PMCID: PMC5880740.

23. Sakowitz S, Bakhtiyar SS, Mallick S, Khoraminejad B, Olmedo M, Croman M, et al. Decreasing rates of colectomy for benign neoplasms: A nationwide analysis. PloS One. 2023;18(10):e0293389. doi:10.1371/journal.pone.0293389 PubMed PMID: 37878628; PubMed Central PMCID: PMC10599571.

24. Ozgur I, Liska D, Cengiz TB, Sapci I, Valente MA, Holubar SD, et al. Colectomy for polyps is associated with high risk for complications and low risk for malignancy: Time for endoluminal surgery? Am J Surg. 2022 Mar;223(3):463–7. doi:10.1016/j.amjsurg.2021.11.030 PubMed PMID: 34906364.

25. Sharma S, McKechnie T, Brennan K, Kuszaj O, Pedroso CM, Doumouras A, et al. Temporal trends in colorectal resections for benign colorectal neoplasia: A retrospective cohort study of the ACS-NSQIP 2016-2022. Colorectal Dis Off J Assoc Coloproctology G B Irel. 2026 Jun;28(6):e70522. doi:10.1111/codi.70522 PubMed PMID: 42286819; PubMed Central PMCID: PMC13263412.

26. Rodrigues R, Geyl S, Albouys J, De Carvalho C, Crespi M, Tabouret T, et al. Effect of implementing a regional referral network on surgical referral rate of benign polyps found during a colorectal cancer screening program: A population-based study. Clin Res Hepatol Gastroenterol. 2021 Mar;45(2):101488. doi:10.1016/j.clinre.2020.06.014 PubMed PMID: 32723672.

27. Alam A, Ma C, Jiang SF, Jensen CD, Webb KH, Boparai ES, et al. Declining Colectomy Rates for Nonmalignant Colorectal Polyps in a Large, Ethnically Diverse, Community-Based Population. Clin Transl Gastroenterol. 2022 May 1;13(5):e00477. doi:10.14309/ctg.0000000000000477 PubMed PMID: 35347095; PubMed Central PMCID: PMC9132519.

28. Ferlitsch M, Hassan C, Bisschops R, Bhandari P, Dinis-Ribeiro M, Risio M, et al. Colorectal polypectomy and endoscopic mucosal resection: European Society of Gastrointestinal Endoscopy (ESGE) Guideline – Update 2024. Endoscopy. 2024 Jul;56(07):516–45. doi:10.1055/a-2304-3219

29. Khan A, Yang H, Habib DRS, Ali D, Wu JY. Development of a machine learning-based tension measurement method in robotic surgery. Surg Endosc. 2025 May;39(5):3422–8. doi:10.1007/s00464-025-11658-9

