## Appendix 2 for "Burden and trends of endoscopically unresectable polyps requiring surgical intervention: A systematic review and meta-analysis"

**PUBMED:**

(

  (

    "Colonic Polyps"[Mesh] OR "Adenomatous Polyps"[Mesh] OR "Colorectal Neoplasms"[Mesh]

    OR "Intestinal Polyps"[Mesh] OR polyp*[tiab] OR adenoma*[tiab]

    OR "laterally spreading"[tiab] OR LNPCP*[tiab] OR "colonic neoplasm*"[tiab]

    OR "colorectal neoplasm*"[tiab] OR "colorectal lesion*"[tiab]

  )

  AND

  (

    colorectal[tiab] OR colon*[tiab] OR rect*[tiab] OR caecal[tiab] OR cecal[tiab]

    OR sigmoid[tiab] OR "ileocecal valve"[tiab] OR "appendiceal orifice"[tiab]

  )

  AND

  (

    (

      "endoscopically unresectable"[tiab] OR "endoscopically non-resectable"[tiab]

      OR "endoscopically nonresectable"[tiab] OR "not amenable to endoscopic"[tiab]

      OR "not suitable for endoscopic"[tiab] OR "unsuitable for endoscopic"[tiab]

      OR "failed endoscopic resection"[tiab] OR "failed endoscopic removal"[tiab]

      OR "endoscopically inaccessible"[tiab] OR "referred for surg*"[tiab]

      OR "surgical referral"[tiab]

      OR "complex polyp"[tiab:~2] OR "complex polyps"[tiab:~2]

      OR "complex lesion"[tiab:~2] OR "complex lesions"[tiab:~2]

      OR "difficult polyp"[tiab:~2] OR "difficult polyps"[tiab:~2]

      OR "challenging polyp"[tiab:~2] OR "challenging polyps"[tiab:~2]

      OR "non-lifting"[tiab] OR nonlifting[tiab] OR "non-lift"[tiab]

    )

    OR

    (

      (

        benign[tiab] OR nonmalignant[tiab] OR "non-malignant"[tiab] OR noncancerous[tiab]

        OR "non-cancerous"[tiab] OR premalignant[tiab] OR "pre-malignant"[tiab]

        OR precancerous[tiab] OR "pre-cancerous"[tiab]

      )

      AND

      (

        "Colectomy"[Mesh] OR colectom*[tiab] OR "segmental resection"[tiab]

        OR "surgical resection"[tiab] OR "bowel resection"[tiab] OR "oncologic resection"[tiab]

      )

      AND

      (

        "Incidence"[Mesh] OR "Registries"[Mesh] OR "Databases, Factual"[Mesh]

        OR incidence[tiab] OR rate[tiab] OR rates[tiab] OR trend*[tiab] OR temporal[tiab]

        OR nationwide[tiab] OR national[tiab] OR "population-based"[tiab]

        OR "population based"[tiab] OR burden[tiab] OR epidemiolog*[tiab]

        OR utilization[tiab] OR utilisation[tiab]

      )

    )

  )

)

NOT

(

  "Case Reports"[pt] OR Editorial[pt] OR Comment[pt]

  OR ("Animals"[Mesh] NOT "Humans"[Mesh])

)

**Scopus:**

TITLE-ABS-KEY ( ( colorectal OR "colorectal polyp*" OR "colonic polyp*" OR "colon polyp*" OR "rectal polyp*" OR "rectosigmoid polyp*" OR "colonic adenoma*" OR "colorectal adenoma*" ) AND ( polyp* OR adenoma* ) AND ( "endoscopically unresectable" OR "endoscopically nonresectable" OR "endoscopically non-resectable" OR "unresectable polyp*" OR "unresectable adenoma*" OR "nonresectable polyp*" OR "non-resectable polyp*" OR "not amenable to endoscopic resection" OR "not suitable for endoscopic resection" OR "unsuitable for endoscopic resection" OR "not amenable to endoscopic removal" OR "not suitable for endoscopic removal" OR "failed endoscopic resection" OR "failed endoscopic removal" OR "referred for surgery" OR "surgical referral" OR ( endoscop* W/3 unresect* ) OR ( endoscop* W/3 nonresect* ) OR ( endoscop* W/3 irresect* ) ) )

**Web of Science:**

TS=(

  polyp*

  OR adenoma*

  OR "colonic polyp*"

  OR "colorectal polyp*"

  OR "adenomatous polyp*"

  OR "laterally spreading"

  OR "laterally spreading tumor"

  OR "laterally spreading tumour"

  OR LST

  OR LNPCP*

)

AND

TS=(

  "endoscopically unresectable"

  OR "endoscopically non-resectable"

  OR "endoscopically nonresectable"

  OR "not amenable to endoscopic"

  OR "not suitable for endoscopic"

  OR "unsuitable for endoscopic"

  OR "not amenable to endoscopic resection"

  OR "not suitable for endoscopic resection"

  OR "failed endoscopic resection"

  OR "failed endoscopic removal"

  OR "referred for surgery"

  OR "surgical referral"

  OR "complex polyp*"

  OR "difficult polyp*"

  OR "challenging polyp*"

  OR "non-lifting"

  OR nonlifting

  OR "non-lift"

  OR nonlift

  OR (endoscop* NEAR/3 unresect*)

  OR (endoscop* NEAR/3 nonresect*)

)

NOT TS=("case report")

**COCHRANE Central:**

#1 polyp*:ti,ab,kw

#2 adenoma*:ti,ab,kw

#3 #1 OR #2

#4 (colorectal NEXT polyp*):ti,ab,kw

#5 (colonic NEXT polyp*):ti,ab,kw

#6 (colon NEXT polyp*):ti,ab,kw

#7 (rectal NEXT polyp*):ti,ab,kw

#8 (rectosigmoid NEXT polyp*):ti,ab,kw

#9 (colorectal NEXT adenoma*):ti,ab,kw

#10 (colonic NEXT adenoma*):ti,ab,kw

#11 #3 OR #4 OR #5 OR #6 OR #7 OR #8 OR #9 OR #10

#12 (endoscopically NEXT unresectable):ti,ab,kw

#13 (endoscopically NEXT nonresectable):ti,ab,kw

#14 (endoscopically NEXT non-resectable):ti,ab,kw

#15 (not NEXT amenable NEXT to NEXT endoscopic NEXT resection):ti,ab,kw

#16 (not NEXT suitable NEXT for NEXT endoscopic NEXT resection):ti,ab,kw

#17 (unsuitable NEXT for NEXT endoscopic NEXT resection):ti,ab,kw

#18 (not NEXT amenable NEXT to NEXT endoscopic NEXT removal):ti,ab,kw

#19 (not NEXT suitable NEXT for NEXT endoscopic NEXT removal):ti,ab,kw

#20 (failed NEXT endoscopic NEXT resection):ti,ab,kw

#21 (failed NEXT endoscopic NEXT removal):ti,ab,kw

#22 (referred NEXT for NEXT surgery):ti,ab,kw

#23 (surgical NEXT referral):ti,ab,kw

#24 (unresectable NEXT polyp*):ti,ab,kw

#25 (unresectable NEXT adenoma*):ti,ab,kw

#26 #12 OR #13 OR #14 OR #15 OR #16 OR #17 OR #18 OR #19 OR #20 OR #21 OR #22 OR #23 OR #24 OR #25

#27 #11 AND #26

**Embase:**

(

'colon polyp'/exp

OR 'adenomatous polyp'/exp

OR 'colorectal neoplasm'/exp

OR polyp*:ti,ab,kw

OR adenoma*:ti,ab,kw

OR 'colorectal polyp*':ti,ab,kw

OR 'colonic polyp*':ti,ab,kw

OR 'colon polyp*':ti,ab,kw

OR 'rectal polyp*':ti,ab,kw

OR 'rectosigmoid polyp*':ti,ab,kw

OR 'colorectal adenoma*':ti,ab,kw

OR 'colonic adenoma*':ti,ab,kw

)

AND

(

'endoscopically unresectable':ti,ab,kw

OR 'endoscopically nonresectable':ti,ab,kw

OR 'endoscopically non-resectable':ti,ab,kw

OR 'not amenable to endoscopic resection':ti,ab,kw

OR 'not suitable for endoscopic resection':ti,ab,kw

OR 'unsuitable for endoscopic resection':ti,ab,kw

OR 'not amenable to endoscopic removal':ti,ab,kw

OR 'not suitable for endoscopic removal':ti,ab,kw

OR 'failed endoscopic resection':ti,ab,kw

OR 'failed endoscopic removal':ti,ab,kw

OR 'referred for surgery':ti,ab,kw

OR 'surgical referral':ti,ab,kw

OR 'unresectable polyp*':ti,ab,kw

OR 'unresectable adenoma*':ti,ab,kw

)
