## Appendix 4 for "Burden and trends of endoscopically unresectable polyps requiring surgical intervention: A systematic review and meta-analysis"

**Table S5. Risk of bias in the 18 included studies: JBI Critical Appraisal Checklist for Prevalence Studies and five supplementary items for administrative and registry data**

| **Study** | **1** | **2** | **3** | **4** | **5** | **6** | **7** | **8** | **9** | **AD1** | **AD2** | **AD3** | **AD4** | **AD5** | **Overall^a^** |
| --- | --- | --- | --- | --- | --- | --- | --- | --- | --- | --- | --- | --- | --- | --- | --- |
| Lee 2013 | Y | Y | Y | Y | Y | Y | Y | U | NA | Y | NA | Y | NA | NR | Low |
| Alam 2022 | Y | Y | Y | Y | Y | Y | U | U | NA | N | Y | Y | P | N | Low |
| Nilsen 2024 | Y | Y | Y | Y | U | U | U | U | NA | Y | Y | N | Y | NR | Moderate |
| Bronzwaer 2018 | Y | Y | Y | Y | N | Y | Y | N | NA | P | NA | N | NA | Y | Moderate |
| Cubiella 2021 | Y | Y | Y | Y | Y | U | N | Y | Y | Y | Y | Y | Y | N | Moderate |
| Paszat 2021 | Y | Y | Y | Y | Y | N | N | U | NA | Y | Y | Y | NA | N | Moderate |
| Rodrigues 2020 | N | Y | N | Y | Y | Y | U | Y | Y | N | NA | N | NA | NR | Moderate |
| Kawaguti 2023 | N | Y | N | Y | Y | Y | Y | NA | NA | N | NA | P | NA | P | Moderate |
| Kilani 2026 | U | U | Y | Y | U | Y | N | U | NA | N | Y | U | N | NR | High |
| Sakowitz 2023 | Y | Y | Y | Y | Y | N | N | N | NA | Y | N | N | N | Y | High |
| Vu 2021 | Y | Y | Y | Y | U | N | U | U | NA | N | Y | U | N | N | High |
| Ozgur 2022 | N | U | Y | Y | Y | Y | N | N | NA | N | Y | N | P | Y | High |
| Sharma 2026 | N | N | Y | Y | U | N | N | U | NA | N | P | N | NA | Y | High |
| Peery 2018 | Y | Y | Y | Y | U | N | N | N | NA | Y | Y | N | P | Y | High |
| Alsakarneh 2026 | N | N | Y | Y | U | N | N | N | NA | N | N | U | N | Y | High |
| Kaltenbach 2025 | N | Y | Y | Y | Y | N | N | U | NA | P | P | N | Y | Y | High |
| Marres 2017 | N | Y | N | Y | Y | Y | N | N | NA | N | NA | P | NA | NR | High |
| Roy 2016 | U | U | Y | U | U | U | U | U | U | N | N | N | N | N | Not assessed^b^ |

Y = yes; N = no; U = unclear; P = partly; NR = not reported; NA = not applicable; — = not assessable.

JBI items: 1, sample frame appropriate; 2, participants sampled appropriately; 3, adequate sample size; 4, subjects and setting described; 5, sufficient coverage of the identified sample; 6, valid methods for identifying the condition; 7, condition measured in a standard, reliable way; 8, appropriate statistical analysis; 9, adequate response rate.

Supplementary items: AD1, representative sampling frame; AD2, diagnostic and procedure codes stated in full; AD3, denominator stable across the observation period; AD4, coding revision within the observation window addressed; AD5, non-elective operations excluded.

^a^ Overall JBI judgement. Ratings for Ozgur 2022, Paszat 2021, Kaltenbach 2025 and Kawaguti 2023 were reached after adjudication of differing domain-level judgements.

^b^ Roy 2016 could not be assessed because of low data availability.

**Table S6. GRADE certainty of evidence, assessed separately within each denominator class**

| **Denominator class** | **Studies (n)** | **Starting certainty^a^** | **Risk of bias** | **Inconsistency** | **Indirectness** | **Imprecision** | **Publication bias^b^** | **Certainty** |
| --- | --- | --- | --- | --- | --- | --- | --- | --- |
| N1; population at risk | 4 | High | Serious (−1) | Very serious (−2) | Serious (−1) | Not serious | Not assessable | Very low |
| N2; all colorectal resections | 9 | High | Serious (−1) | Very serious (−2) | Serious (−1) | Not serious | Not assessable | Very low |
| N3; all polyps or complex polyps detected | 9 | High | Serious (−1) | Very serious (−2) | Serious (−1) | Not serious | Not assessable | Very low |
| N4; all colonoscopies or screening episodes | 6 | High | Serious (−1) | Serious (−1) | Serious (−1) | Not serious | Not assessable | Low |

^a^ Assessment began at high certainty, following the GRADE approach for questions of event rate or baseline risk (Iorio et al., 2015), in which observational studies are not automatically downgraded.

^b^ Publication bias could not be assessed in any class: no class contained ten or more independent data sources, the minimum generally regarded as necessary for meaningful assessment of funnel plot asymmetry.

Principal driver of downgrading, by class: N1, inconsistency in the direction of effect, compounded by fixed population denominators in both anchor studies; N2, heterogeneity of denominator definition within a single nominal class; N3, heterogeneity of numerator and lesion entry criteria; N4, risk of bias and indirectness, inconsistency being less severe than in the other three classes.
